# The Utility of Ultra-Deep RNA sequencing in Mendelian Disorder Diagnostics

**DOI:** 10.1101/2025.01.28.25321295

**Authors:** Sen Zhao, Jefferson C. Sinson, Shenglan Li, Jill A. Rosenfeld, Gladys Zapata, Kristina Macakova, Mezthly Pena, Becky Maywald, Kim C. Worley, Lindsay Burrage, Monika Weisz Hubshman, Shamika Ketkar, William Craigen, Lisa Emrick, Undiagnosed Diseases Network, Tyson Clark, Gila Yanai Lithwick, Zohar Shipony, Christine Eng, Brendan Lee, Pengfei Liu

## Abstract

Clinical RNA-seq has become an essential tool for resolving variants of uncertain significance (VUS), particularly those affecting gene expression and splicing. However, most reference data and diagnostic protocols employ relatively modest sequencing depths (∼50-150 million reads), which may fail to capture low-abundance transcripts and rare splicing events critical for accurate diagnoses. We evaluated the diagnostic and translational utility of ultra-high-depth (up to ∼one billion unique reads) RNA-seq in four clinically accessible tissues (blood, fibroblast, LCL, and iPSC) using Ultima sequencing platform. After validating the performance of Ultima RNA-seq, we investigated how increasing depth affects gene and isoform detection, splicing variant discovery, and clinical interpretation of VUS. Deep RNA-seq substantially improved sensitivity for detecting lowly-expressed genes and isoforms. At ∼1 billion reads, near-saturation was achieved for gene-level detection, although isoform-level coverage continued to benefit from even deeper sequencing. In two clinical cases with VUS, pathogenic splicing abnormalities were undetected at ∼50 million reads but emerged at 200 million reads, becoming even more pronounced at ∼one billion reads. Using deep RNA-seq data, we constructed a novel resource, MRSD-deep, to estimate the minimum required sequencing depth to achieve desired coverage thresholds. MRSD-deep provided gene- and junction-level guidelines, aiding labs in selecting suitable coverage targets for specific applications. Leveraging deep RNA-seq data on fibroblast, we also built an expanded splicing-variation reference that successfully identified rare splicing events missed by standard-depth data. Our findings underscore the diagnostic and research benefits of deep RNA-seq for Mendelian disease investigations. By capturing rare transcripts and splicing events, ultra-high-depth RNA-seq can facilitate more definitive variant interpretations and enrich splicing-reference databases. We anticipate that cost-effective deep sequencing technologies and robust reference cohorts will further advance RNA-based diagnostics in precision medicine.

## Introduction

Sequencing depth in next-generation sequencing (NGS) refers to the number of reads covering a specific region or transcript. It is typically calculated on a genome- or transcriptome-wide scale to describe the overall characteristics of the sequenced sample in two key aspects: (1) the proportion of targeted regions adequately interrogated for qualitative evaluation, and (2) the magnitude of alterations that can be quantitively assessed at the covered targets.

In DNA sequencing, sequencing depth is often straightforward to define because the human nuclear genome has a uniform copy number of two across most regions. Consequently, depth is conveniently measured as the average number of times each base is sequenced, making it a reliable indicator of coverage. Extensive studies have demonstrated how varying sequencing depths impact variant detection ^1^, leading to clear recommendations for minimum depth requirements in professional guidelines ^2,3^.

In contrast, RNA sequencing (RNA-seq) presents unique challenges in defining and interpreting sequencing depth due to the heterogeneity of transcript expression levels. Unlike DNA, where coverage is relatively uniform, RNA-seq targets range from highly expressed transcripts to rare, low-abundance transcripts. As a result, sequencing depth in transcriptomics is typically defined by the total number of mapped reads, rather than the average fold coverage per base.

The currently recommended sequencing depths for RNA-seq in human cells are based on a combination of early experimental insights, subsequent optimization, and cost considerations. Early studies, such as those using RNA standards from the External RNA Control Consortium, highlighted the challenge of detecting low-abundance transcripts at ∼12.4 million mapped reads^4^. Saturation studies focusing on differential expression analysis demonstrate that ∼36 million mapped reads are sufficient to detect genes with high expression levels, whereas accurate quantification of low-expression genes requires sequencing depths of up to 80 million reads ^5^. These foundational studies have informed best practices for RNA-seq, but they were largely developed in research settings and do not fully address the unique challenges of clinical RNA sequencing, particularly the reliance on clinically accessible tissues (CAT).

In clinical diagnostic RNA-seq, the use of CATs, such as blood or fibroblasts, is necessary due to the inaccessibility of disease-relevant tissues. While this approach has improved diagnostic yield for Mendelian disorders by aiding the interpretation of variants of unknown significance (VUS) when combined with DNA sequencing ^6–8^, it presents inherent limitations.

For instance, a recent study found that 40.2% of all genes were inadequately represented by at least one CAT, while 6.3% were inadequately represented across all CATs, even at a depth of ∼50 million total reads ^9^. This limitation contrasts with research applications of RNA-seq, where sequencing is typically conducted on proxy tissues or cell lines that closely resemble the affected tissue, allowing for a more direct investigation of pathogenic mechanisms.

Although the range of 50 to 150 million reads for human transcriptome sequencing has been commonly adopted in clinical RNA-seq practices, this range has largely been driven by practical considerations, including historical use in research, technical feasibility, and cost.

However, there has been limited emphasis on systematically evaluating deeper sequencing depths to assess their potential in improving diagnostic utility, particularly when using CATs.

Here, we aim to evaluate the diagnostic and translational utilities of ultra-high-depth RNA-seq (deep RNA-seq) in clinical diagnostics. Using mostly natural sequencing-by-synthesis from Ultima Genomics, a cost-effective platform, we sequenced four CATs, i.e., blood, fibroblast, lymphoblastoid cell lines (LCL), and induced pluripotent stem cells (iPSC), at depths exceeding one billion unique reads. We assessed how increased sequencing depth impacts the detection of genes and isoforms, providing case examples where molecular diagnoses were validated through deep RNA-seq. Furthermore, we leveraged these deep-RNA-seq datasets to create resources for genetic diagnoses.

## Results

### Technical Evaluation of Ultima RNA-seq

To assess the reliability of Ultima RNA-seq for transcriptome profiling, we first compared its performance metrics against Illumina RNA-seq using 15 fibroblast samples that were sequenced with both technologies (**Table S1**). Each pair of Ultima/Illumina data was downsampled to the same total read counts to ensure a fair comparison. Both platforms achieved similar mapping rates and exonic rates (**Figure S1a**), indicating comparable data quality in aligning reads to the transcriptome. Notably, Ultima sequencing produced longer read lengths, resulting in a higher split rate (**Figure S1b**) and the detection of a greater number of splicing junctions (**Figure S1c**). Furthermore, the transcriptomic expression profiles generated by Ultima and Illumina RNA-seq are well-correlated (**Figure S1d and Figure S2**), supporting the comparability of the two platforms for gene quantification.

We then benchmarked the performance of Ultima RNA-seq using the ‘gold standard’ RNA-seq reference established in our recent study ^10^. The reference was generated from publicly available short-read and long-read data for the Genome in a Bottle (GIAB) sample GM24385 (HG002). Positive and negative gene/junction sets were used to assess the sensitivity and specificity of Ultima sequencing. The GM24385 sample was sequenced at varying depths, ranging from 39 million to 826 million reads (**Table S2**). Gene and junction detection sensitivity in Ultima RNA-seq was comparable to Illumina at similar sequencing depths and further improved with deeper sequencing (**Table S2**). While specificity decreased at higher sequencing depths, it is important to note that the observed ‘false positive’ calls could represent true positives that were not detected in our ‘gold standard’ reference, which was derived from standard-depth RNA-seq data.

Taken together, these data demonstrate that Ultima RNA-seq delivers reliable and accurate gene and junction detection, with performance comparable to Illumina RNA-seq. Importantly, the data suggest that Ultima RNA-seq has the potential to achieve even greater accuracy with higher sequencing depths, making it a promising technology for transcriptome analysis.

### Deep RNA-seq on clinically accessible tissues

After validating the sequencing platform, we applied Ultima RNA-seq to four CATs (blood, fibroblast, lymphocyte, and iPSC; n = 3 for each CAT). Our goal was to explore the impact of ultra-high sequencing depth on transcript detection in tissues commonly used for clinical testing. Each sample was sequenced to a target depth of 2 billion (2000M) total reads (**Table S1**), with unique molecular identifier (UMI)-based library designs employed to evaluate duplication rates (**Figure S3**). After mapping and UMI-based deduplication, the number of unique reads ranged from 1281M to 1726M (**Table S3**), and the corrected duplication rates hovered around 20% (**Table S3**), indicating an adequate library complexity.

To evaluate whether *in silico* downsampling would faithfully represent different sequencing depths, we sequenced three samples at a range of depths *in vitro* and compared their gene expression profiles to *in silico* downsampled data. The two sets of data showed high concordance, validating our simulation approach (**Table S4**). We further confirmed that high-depth RNA-seq generated high-quality reads, as indicated by stable exonic rates even at higher depths (**Figure S4**).

We next investigated how increasing sequencing depth affects gene detection (**Data S1**). For multi-exon genes, we defined “detection” as > 50 total reads with > 2 junction-spanning reads. Single-exon genes required > 100 total reads. These thresholds were chosen based on junction ratios of genes at different read counts and manual inspection of the raw data (**Figure S5**).

Overall, iPSC yielded the highest number of detected genes among the four CATs (**Figure 1a**), consistent with previous findings that iPSCs can express a wide variety of genes ^11^. LCL detection performance was modest at lower depths but converged with that of blood and fibroblast at higher depths – likely due to the larger number of low-expressing genes in LCL (**Figure S6**). Across all four CATs, each additional million reads uncovered 10-30 new genes at 100 million reads. At 1000M (one billion) reads, the detection rate slowed, reaching 1-2 new genes per million reads (**Figure 1a**), suggesting a saturation effect for gene detection beyond this point. When we focused on OMIM genes (n=4875), 3908 to 4332 of them are reliably detected at 1000M reads in the four CATs (**Figure 1a, Data S1**).

**Figure 1.**
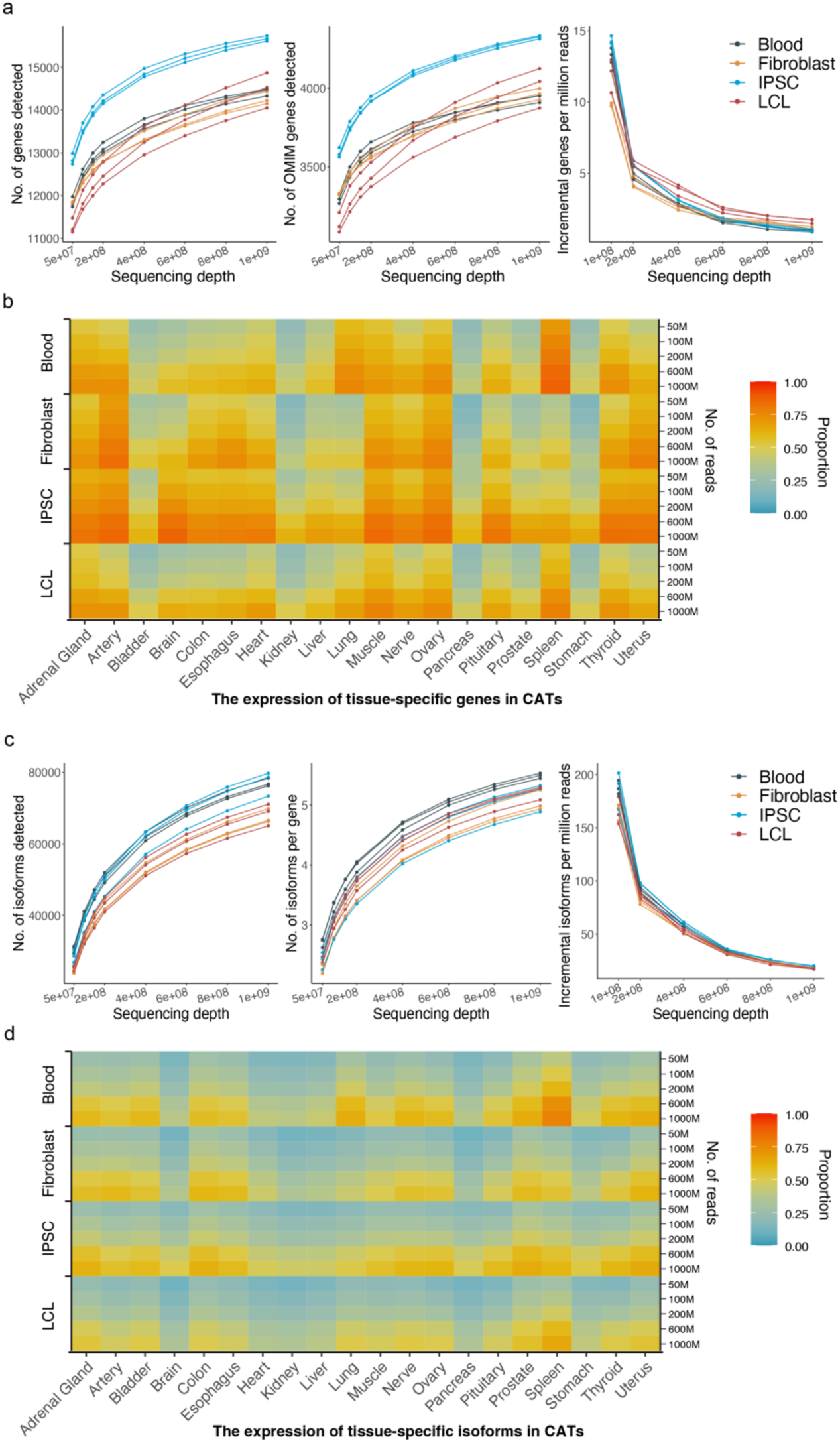
Transcriptome sequencing at ultra-high depth enables detection of higher number of genes and isoforms in clinically accessible tissues. a, Relationship between transcriptome sequencing depth and the number of genes detected (out of 19217 coding genes), the number of OMIM genes detected (out of 4875 genes), and incremental genes detected per million reads. b, Proportion of tissue-specific genes detected in clinically accessible tissues (CATs) across different sequencing depths. c, Relationship between transcriptome sequencing depth and the number of isoforms detected, the average number of isoforms detected per gene, and incremental isoforms detected per million reads. d, Proportion of tissue-specific isoforms detected in CATs across different sequencing depths. Tissue-specific genes and isoforms were defined based on data from the GTEx database.

We further explored whether deep sequencing could help overcome tissue-specific gene expression gaps. Interestingly, many genes with low expression in CATs (TPM < 0.1) - a group likely to benefit from deep sequencing - were found to have higher expression levels (TPM > 1) in at least one non-CAT tissue (**Figure S7**). Building on this, we hypothesized that tissue-specific genes might particularly benefit from higher sequencing depths. To test this, we identified tissue-specific genes (tissue preference score > 2) using data from GTEx. As expected, these genes showed depleted expression in standard depth (∼100M) GTEx CAT data (fibroblast, blood, and LCL) (**Figure S8**). However, sequencing at ∼1000M unique reads detected 61-75% of these tissue-specific genes in our CATs (**Figure 1b**, **Data S2**) - a significant improvement over 50M reads, which captured only 40-50% (**Figure 1b, Data S2**). These results suggest that deep RNA-seq can substantially enhance the utility of CATs as proxies for disease tissues.

We further examined the detection of transcript isoforms. Around 60,000 to 80,000 isoforms were detected in each CAT at 1000M reads, corresponding to 4-6 isoforms per gene (**Figure 1c, Data S3**). Consistent with its broad expression profile, iPSC yielded the most isoforms in absolute terms, whereas blood exhibited the highest number of isoforms per gene-likely reflecting its cellular heterogeneity (**Figure 1c**). The addition of each million reads revealed 100-200 previously undetected isoforms at 100M reads but only 15-20 at 1000M reads (**Figure 1c**), indicating saturation at the isoform level. Focusing on tissue-specific isoforms (tissue preference score > 2), we found that 44-55% were captured at 1000M reads (**Figure 1d, Data S4**) – over two-fold more than at 50M reads. Still, isoform detection remained modest for tissues such as the brain and liver that undergo particularly abundant alternative splicing (**Figure 1d, Data S4**).

Finally, we explored the potential of using deep RNA-seq data to inform the design of depletion assays aimed at improving the detection of underrepresented genes. Experimental knockdown of high-expressing genes prior to sequencing has been proposed as a strategy to free reads for lower-expressed genes, thereby enhancing their detection ^12^. Leveraging our deep RNA-seq data, we computationally simulated this approach to evaluate its potential enrichment effects. At standard 100M read coverage, in silico removal of 3000 genes increased the detection of an additional ∼800 (LCL) to ∼2000 (iPSC) low-expression genes, with saturation reached at these levels (**Figure S9**). Using deep RNA-seq data, which better represents low-expression genes, we found that removing ∼7500 genes allowed for an additional ∼ 3000 (LCL) to 3700 (iPSC) detections (**Figure S9**). These findings offer valuable insights for optimizing gene knockdown strategies and experimental designs to enrich transcriptome coverage for low-expression genes.

### Deep RNA-seq enables the diagnosis of Mendelian disorders

To investigate the clinical relevance of high-depth RNA-seq, we selected two fibroblast samples originating from patients whose DNA sequencing had returned only variants of uncertain significance (VUS). Previous RNA-seq at standard depth (∼50M) had not detected any clinically relevant abnormal RNA-level changes. By re-analyzing each sample under deep RNA-seq, we identified disease-relevant findings in both cases.

Case #1 involved an intronic variant c.5980-17G>A in *ITPR1*, classified as a VUS based on DNA data alone ^13^. Because *ITPR1* is only weakly expressed in fibroblasts, standard depth RNA-seq at 50M reads showed no significant expression or splicing abnormalities (**Figure 2a**). However, at 200M reads, we observed reads supporting a cryptic splice site (P = 0.009, Fraser, **Figure 2a**). At 1000M reads, the cryptic splice site became clearer (P = 0.002, Fraser, **Figure 2a**) and a significant downregulation was detected (P = 0.04, fold change = 0.55, Outrider). This direct measurement of splicing consequences prompted an upgrade of the variant to likely pathogenic. Of note, while *ITPR1* expression can be upregulated through cell trans-differentiation ^14^ (**Figure 2a**) or CRISPR activation ^15^, these methods significantly prolong turnaround times. By contrast, deep RNA-seq can more rapidly illuminate splicing defects *in situ*.

**Figure 2.**
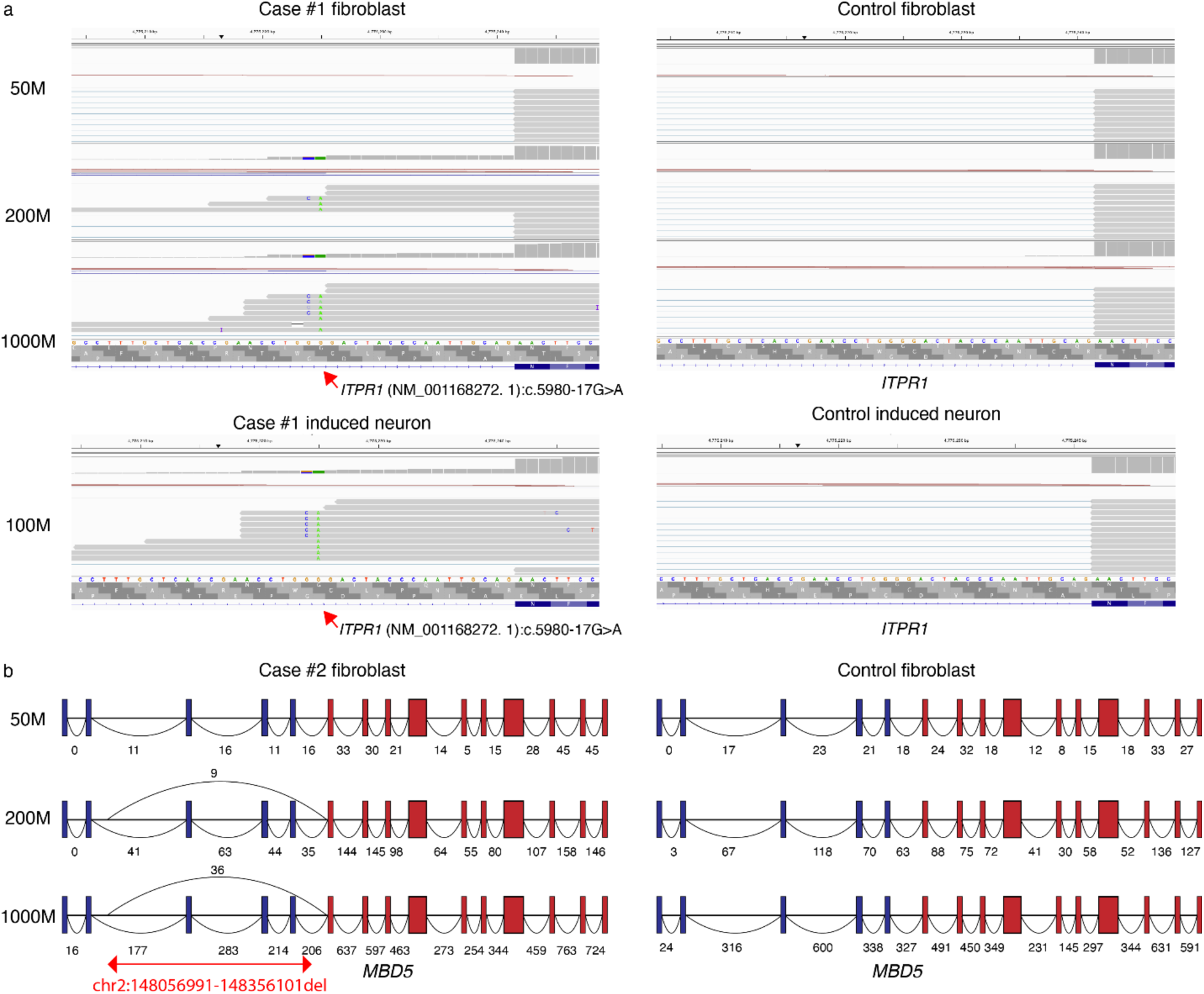
Deep RNA sequencing identifies sparsely expressed abnormal transcripts in clinically accessible tissues, aiding molecular diagnosis. a, RNA-seq data from a patient carrying a heterozygous c.5980-17G>A variant in *ITPR1* (indicated by the red arrow) revealed a cryptic splice site. Sequence alignments at the variant site were shown for fibroblasts or induced neuron cells from the case and control at different sequencing depths. b, An abnormal junction spanning multiple exons in *MBD5* caused by a DNA deletion (red arrows) was not detected at lower sequencing depths but becomes evident as sequencing depth increases. Exons in blue (noncoding) and exons in red (coding) represent two short transcripts, which together form a full-length transcript enriched in neural tissues.

Case #2 featured a deletion involving exons 3-5 and their flanking introns in *MBD5*, which is commonly expressed in two short transcripts (exons 1-5 [noncoding] and exons 6-15 [coding]) and one full-length transcript (**Figure 2b**). The full-length isoform is most relevant in the interpretation of this variant, but it has low expression in fibroblast. At 50M reads, we detected no abnormal junction reads. Increasing the depth to 200M revealed nine junction reads bridging intron 2 and exon 6, and raising to 1000M uncovered 36 such reads. Deep RNA-seq thus enabled us to detect low-abundance isoforms carrying disease-causing splicing events.

### MRSD-deep: a resource for determining the optimal depth for clinical RNA-seq

While these case studies confirm the clinical utility of high sequencing depth, a practical question remains: how can clinicians systematically determine the optimal depth for a given set of candidate genes? A previous study by Rowlands et al. proposed the concept of minimum required sequencing depth (MRSD), defining the total sequencing reads needed so that a desired proportion of splice junctions exceeds a coverage threshold ^16^. The MRSD calculations were based regular-depth GTEx data, making many genes “unfeasible” due to low expression levels ^16^. We therefore computed a new set of MRSD values, termed MRSD-deep, leveraging our Ultima-based data in which each CAT was sequenced to ∼1000M unique reads.

First, we calculated MRSD for canonical junctions of each GENCODE multi-exon gene, using the median junction read count from our three deep RNA-seq datasets per CAT. A junction was deemed “unfeasible” if the median junction read count was zero. We computed MRSD across multiple desired junction read thresholds and junction proportions (**Figure 3a**). As expected, higher coverage thresholds lead to higher MRSD values (**Figure 3a**), and a higher proportion of junctions required to be covered per gene increased the likelihood of certain genes being marked unfeasible (**Figure 3b**). Our MRSD-deep values were generally lower than those reported in the original study (**Figure 3c, Figure S10**), likely reflecting differences in read length and split rates between Ultima and standard Illumina platforms. Moreover, our approach yielded approximately 100% more feasible genes (**Figure 3c)**, largely due to improved sensitivity for low-expressing genes in CATs.

**Figure 3.**
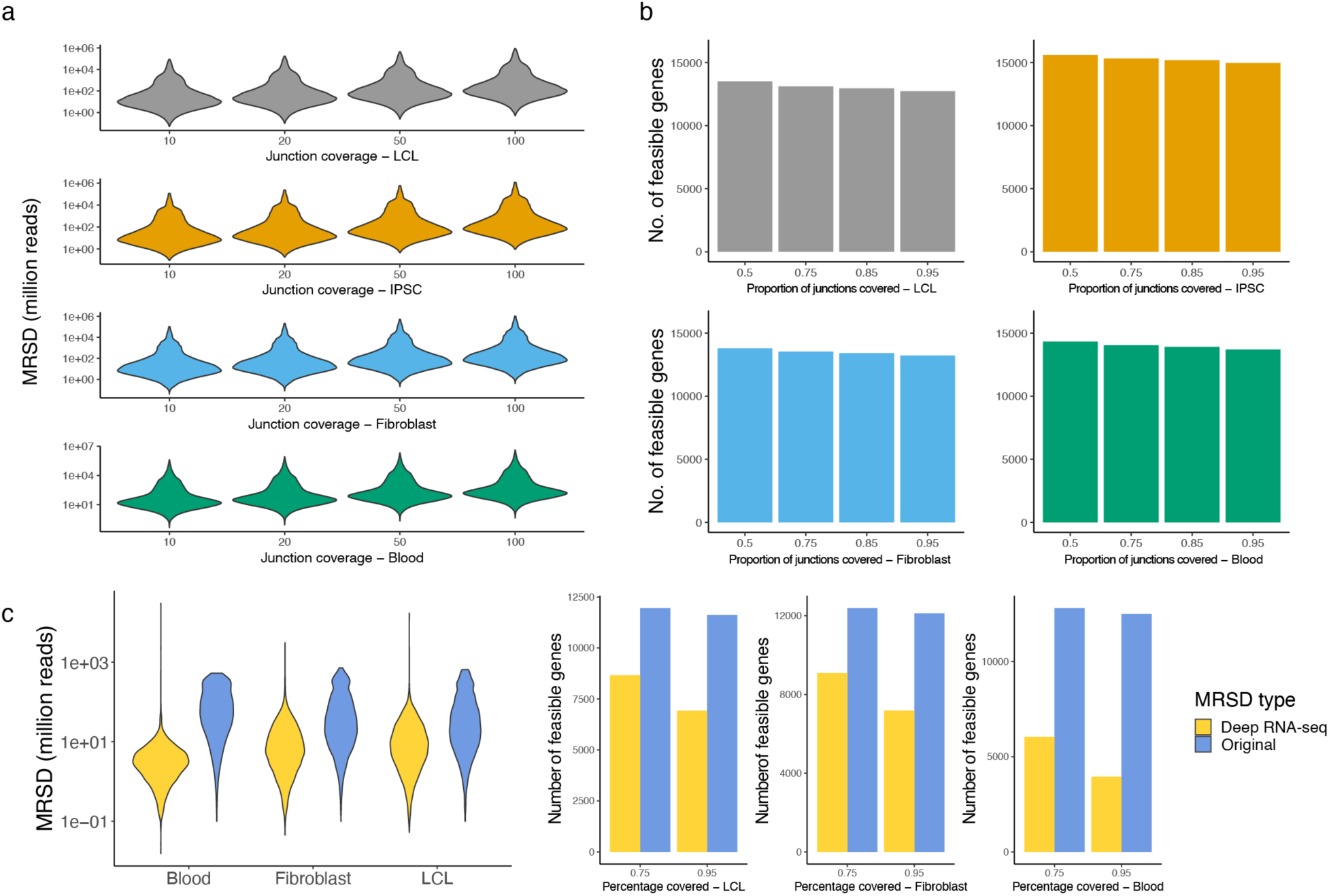
Minimum required depth analysis using deep RNA-seq. a, Junction-level minimum required sequencing depth (MRSD), the total sequencing depth required for 95% of splice junctions for a candidate gene to be covered at the desired number of reads. b, The number of genes feasible for MRSD analysis in four clinically accessible tissues. A gene is only feasible when a given proportion of all splicing junctions have non-zero median depth in our deep RNA-seq data. Therefore, when a larger proportion of junctions are required to be covered, the number of feasible genes decreases. c, Comparison of MRSD metrics between our data and the original MRSD data. The distribution of MRSD was calculated at a required junction coverage of 95% and junction read of 10. The number of feasible genes was calculated based on a total of 16579 genes that are available in the original MRSD.

In addition to splice junction coverage, accurate detection of gene expression alterations is also vital in a molecular diagnostic setting. Generally, one uses a control reference set to assess whether the gene’s expression is aberrant. The required read count for robust calls depends on the dispersion parameters of the candidate gene in that reference. For example, based on a fibroblast reference dataset of 73 RNA-seq datasets ^10^, a gene like *ITPR1* (theta = 13.19) requires ∼5000 reads to reliably detect a fold change of 80%, whereas *MBD5* (theta = 120.47) requires ∼50,000 (**Figure 4a, Figure S11**). Indeed, the average read count of *ITPR1* in fibroblast deep RNA-seq is 5505.17, enabling the detection of its expression change (P = 0.04, fold change = 0.55, Outrider).

**Figure 4.**
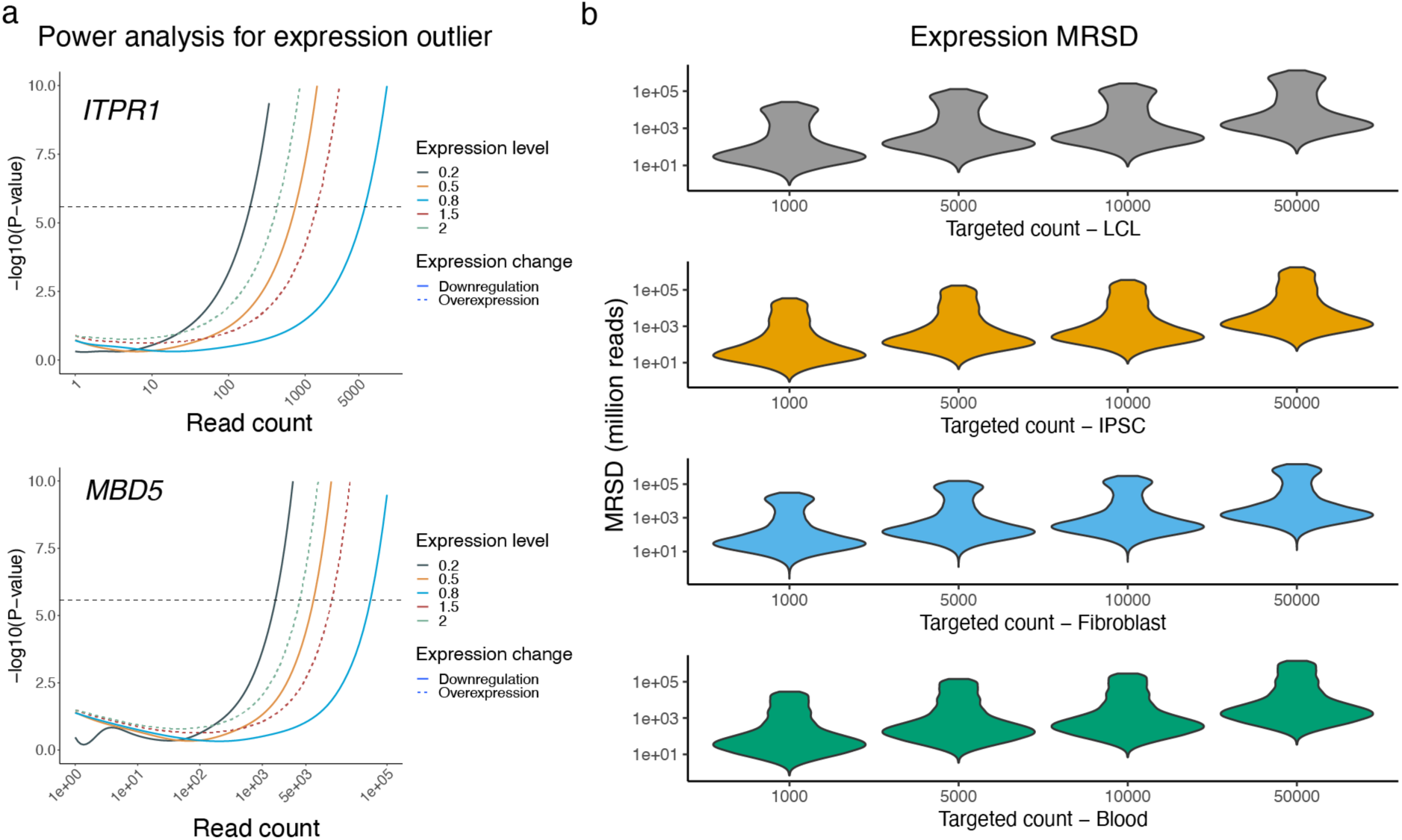
Minimum required depth analysis at expression level. a, Power analysis of expression outlier analysis. For different genes, the required read counts to enable the detection of expression outlier depend on their expression variations in the reference panel. b, Expression-level MRSD, the total sequencing depth required for a candidate gene to be covered at the desired number of reads.

Unlike the original MRSD work, which focused solely on splice junction coverage, our analysis extends the MRSD framework to account for gene expression reads (**Figure 4b**), introducing a novel dimension to the method. Both LCL and fibroblast presented a bimodal MRSD distribution, consistent with our earlier finding that these tissues harbor a large number of genes with low expression (**Figure S6**). Taken together, these MRSD-deep values can help clinicians decide on the sequencing depth needed for a given diagnostic target. **Supplementary Data S5 and S6** provide full MRSD-deep metrics for splice junctions and gene-level expression, respectively.

### Naturally occurring splicing variations

Finally, beyond direct diagnostics, deep RNA-seq from diverse populations can illuminate the spectrum of alternative splicing events. Naturally occurring alternative splicing in the population can inform diagnostics, as these events may become the dominant splicing patterns when canonical splicing is disrupted ^8^. This information is highly valuable for predicting the consequences of mutations that affect canonical splicing sites ^17,18^, as well as for mapping trait-associated splicing variations ^19^. However, these foundational concepts have primarily been demonstrated using reference datasets like GTEx, typically generated at ∼100M reads, which may miss splicing events of very low abundance.

To address this gap, we generated deep RNA-seq data for an additional 15 fibroblast samples at ∼one billion unique reads each, thus collecting 18 fibroblast deep RNA-seq in total. We also collected 18 fibroblast RNA-seq datasets at ∼100M reads from our recent study to serve as a “standard-depth” control ^10^. Using an approach similar to SpliceVault ^18^, we constructed a new splicing variation database from these data. We ranked each detected alternative splicing event by its prevalence in the 18 reference samples (**Data S7**). Because pathogenic variants often co-opt low-frequency alternative splicing events, high-depth RNA-seq that reveals these events could predict pathogenic outcomes more accurately.

For example, an intronic disease-causing variant in *TMEM161B* that leads to exon 8 skipping was confirmed in the patient sample by standard-depth RNA-seq (**Figure 5a**). The same exon-skipping event was detectable at low abundance from healthy control samples only at 1000M reads but not at 100M (**Figure 5a**). Another disease-associated intronic variant in *RPL13* caused a cryptic donor site. While this cryptic donor site was observable at 100M reads in control samples, substantially more supporting reads were detected at 1000M (**Figure 5b**). These examples illustrate how deep RNA-seq enables the detection of rare pathogenic splicing events that might otherwise remain undetected at standard sequencing depths.

**Figure 5.**
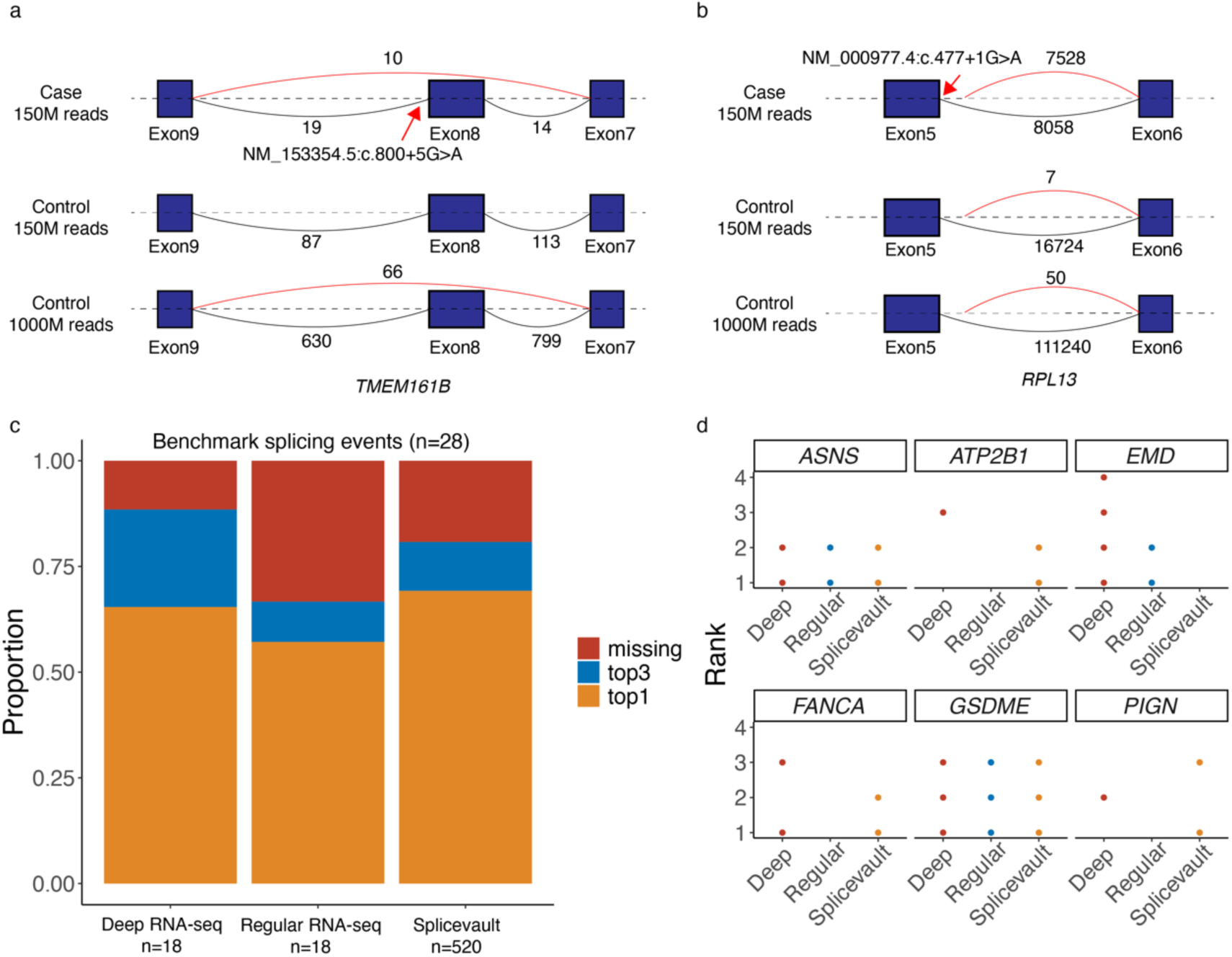
The utility of naturally occurring splicing variation in diagnostics. a, An example of a case with exon skipping (red arch) of *TMEM161B* caused by an intronic variant (red arrow). The junction plots of the case and a control sample sequenced at different depths are shown. b, Another example of a case with cryptic splice site (red arch) of *RPL13* caused by an intronic variant (red arrow). The junction plots of the case and a control sample sequenced at different depths are shown. c, The performance of the deep RNA-seq, regular RNA-seq, and the original SpliceVault in predicting the splicing consequences of 28 known positive splicing variants. d, The performance of the deep RNA-seq, regular RNA-seq, and the original SpliceVault in predicting six variants with multiple splicing consequences.

To benchmark this approach, we compiled 28 disease-causing variants with known splicing outcomes from our inhouse database and a previous study (**Data S8**) ^18^. We compared the rank of each disease variant’s main splicing consequence in three datasets: (i) our deep RNA-seq database, (ii) our regular-depth control, and (iii) the original SpliceVault (n = 520 samples at ∼100M reads). Our deep RNA-seq database outperformed SpliceVault in detecting the pathogenic outcomes within the top-3 rank, though it achieved a slightly lower top-1 rank (**Figure 5c**). In contrast, the lower sequencing depth in the 18 standard-depth samples significantly hindered their ability to capture disease-causing splicing outcomes. For the eight variants with multiple confirmed splicing consequences, deep RNA-seq and SpliceVault each captured different subsets, highlighting their complementarity (**Figure 5d).**

In short, these results underscore how deep RNA-seq can substantially enhance public splicing-reference datasets, even with relatively small sample sizes. By mapping rare splicing events, deep RNA-seq improves our ability to predict the effects of pathogenic variants, ultimately strengthening diagnostic and research capabilities.

## Discussion

In this study, we systematically evaluated how ultra-high-depth RNA-seq can enhance diagnostic yield and facilitate the interpretation of variants of uncertain significance. By sequencing CATs at depths up to one billion reads, we uncovered transcripts and isoforms that typically remain undetected at standard depth (∼50 - 150M). Our results revealed a near-saturation point for gene detection at around 1000M reads, although isoform detection continued to benefit from deeper sequencing. Importantly, we showed that ultra-high-depth RNA-seq not only improves characterization of challenging, lowly-expressed genes in CATs, but can also illuminate rare splicing events with potential diagnostic relevance.

To address practical needs in clinical testing, we developed MRSD-deep matrices for both splice junction and gene expression coverage. Clinicians and researchers can consult these resources to select depth thresholds tailored to specific genes of interest. For highly expressed genes, standard depths might suffice; for low-expression genes with complex isoforms, deeper coverage (e.g., >1000M reads) may be necessary. In cases where MRSD calculations suggest impractical depth requirements, alternative approaches such as target enrichment assays (e.g., panel-based RNA-seq) ^20^, tissue-specific sampling, cell transdifferentiation ^14^, or transactivation ^15,21^ may be considered.

Beyond its direct diagnostic applications, deep RNA-seq enriches splicing-reference databases by capturing alternative splicing events that occur at extremely low abundance. Previous studies focused on expanding the sample sizes to discover splicing variations ^8,18^. Deep RNA-seq provided another dimension of information by capturing splicing variations at extremely low abundance. By integrating both depth and sample size, researchers can move closer to a comprehensive catalog of naturally occurring splicing patterns essential for accurately predicting the impact of intronic and regulatory variants.

While short-read data reveal abundant isoforms and splicing events, some complex transcript structures might remain unresolved. Future investigations could combine deep short-read sequencing with targeted long-read approaches to validate isoform architectures and confirm splicing junctions in clinically actionable genes. Our study shows how high-depth sequencing benefits from well-curated control datasets. As more researchers adopt deep RNA-seq, building larger reference cohorts (stratified by tissue type and ancestry) will further improve expression and splicing outlier detection for rare diseases.

In conclusion, our evidence-based investigation highlights the clinical diagnostic and translational advantage of sequencing the transcriptome at ultra-high depths. By uncovering low-abundance genes and isoforms, elucidating rare splicing variants, and guiding practical decisions regarding sequencing coverage, deep RNA-seq represents a powerful adjunct to current genetic testing paradigms. We anticipate that future improvements in sequencing technologies and collaborative reference databases will further expand its utility in medical genetics.

## Methods

### Ethics approval

This study was approved by the Institutional Review Board (IRB) at Baylor College of Medicine (H-42680). The study subjects were originally recruited through informed consent approved by the IRB at the National Human Genome Research Institute (15HG0130) or BCM (H-34433 and H-44172), and then de-identified for the current study.

### Sample collection

A total of 42 samples were included in this study, including 33 fibroblast samples, 3 blood samples, 3 lymphoblastoid cells (LCL), and 3 PBMC-derived pluripotent stem cells (iPSC). Among them, 12 samples (3 for each sample type) were used in the main sequencing depth analysis, 15 fibroblast samples were used for validation of the Ultima platform, and other 15 fibroblast samples were used to construct the splicing variation dataset.

Samples were collected from three sources:

1. Three LCL samples (GM24385, GM12878 and GM12877) and one IPSC sample (GM27730) were obtained from the Coriell Institute.
2. All fibroblast samples and one blood sample were obtained from the Undiagnosed Diseases Network (UDN).
3. Three blood samples and two IPSC samples were collected from healthy volunteers.

A complete list of samples and their demographic information is provided in Table S1.

### RNA extraction

For fibroblasts, LCL, and IPSC, RNA was extracted from around 1×10^7^ cells using the RNeasy mini kit (Qiagen) following the manufacturer’s instructions with the inclusion of an on-column gDNA removal step. For blood samples, RNA was extracted using QIAamp RNA blood mini kit. The integrity and quality of the RNA were assessed using the Qubit 4 Fluorometer and the Qubit RNA HS Assay Kit (ThermoFisher) and was required to reach the following criteria:

1. Amount ≥ 1.5 µg, Volume: ≥ 20µL
2. A260/280: 1.7 – 2.2
3. RIN ≥ 6.0

### Illumina RNA-seq

We sequenced 15 fibroblast samples on both Illumina and Ultima platforms. RNA-seq library was prepared using the Illumina Stranded mRNA prep kit. Sequencing was conducted on the Illumina HiSeq 2500 platform which produced pared-end short read data at 150bp. The fibroblast samples were sequenced to a target depth of 50 million reads.

### Ultima RNA-seq

The UG100 machine was used for Ultima sequencing. A standard Ultima mRNA library (Figure S3) was prepared for the 15 fibroblast validation samples using NEBNext® Ultra™ II Directional RNA Library Prep with Sample Purification Beads in conjunction with NEBNext® Poly(A) mRNA Magnetic Isolation Module. For the 15 fibroblast samples that were also sequenced on the Illumina platform, their Ultima libraries were sequenced at a target depth of 100 million reads. Raw fastq data from Ultima was downsampled to the same read number of the paired Illumina data using seqtk v1.4.

For deep RNA-seq, an additional 10-bp unique molecular identifier (UMI, 5-bp at both ends) was added to the standard Ultima library design (Figure S3), which allowed the calculation of the duplication rate. For sequencing depth analysis, we performed deep RNA-seq at a target depth of 2 billion reads on 12 samples, including 3 fibroblast samples, 3 blood samples, 3 IPSC samples, and 3 LCL samples. After UMI-based dedup, in silico downsampling was performed on each data to simulate RNA-seq at different total reads (50M, 100M, 150M, 200M, 400M, 600M, 800M, and 1000M). To verify the accuracy of this in silico approach, we carried out multi-depth sequencing (at targeted depths of 50M, 100M, 200M, 400M, 800M, and 2400M total reads) on three samples (GM24385 [LCL], UD1902P0001 [fibroblast], and BG2111P0002 [IPSC]) and compared their gene expression profiles with those generated from the corresponding simulated data. We also performed deep RNA-seq at a target depth of 2 billion reads on an additional 15 fibroblast samples from the UDN. These deep RNA-seq data, together with the three other fibroblast deep RNA-seq data mentioned above, were used to construct the alternative splicing resource.

### RNA-seq data analysis

Raw fastq data from Illumina or Ultima sequencer were processed using the same pipeline adapted from the Genotype-Tissue Expression (GTEx) version 10 pipeline (https://github.com/broadinstitute/gtex-pipeline/blob/master/rnaseq/README.md). Sequencing FASTQ data were aligned to the reference genome GRCh38 with STAR v2.7.8a_sentieon and SAMtools 1.15.1/HTSlibv1.10.2. Picard v2.23.3 was used to mark duplicates. Gene expressions were quantified by RNA-SeQC v2.4.2. Isoform-level quantification was calculated by RSEM v1.3.3. Transcripts were annotated with GENCODE v39. FastQC v0.11.9 was used to generate quality control measurements.

For Ultima data with UMI, cutadapt v3.4 was used to look for adaptor sequences, and umi_tools v 1.1.4 was used to extract UMI sequences and perform UMI-based deduplication. High-expressing genes (read count > 10000) were excluded when calculating the duplication rate because a ten-base UMI can overestimate the duplication rate for these genes.

In the sequencing depth analysis, a gene was defined as expressed if 1) the gene is a multi-exon gene with a read count (from RNA-SeQC) of more than 50 and junction reads (from STAR) of more than two or 2) the gene is a single-exon gene with a read count of more than 100. Similarly, an isoform was defined as expressed if the expected count of the isoform (from RSEM) is more than 50.

### RNA-seq Benchmark data

The GM24385 (HG002) cell line is a human lymphoblastoid cell line that originated from a female donor and is commonly used for benchmarking and validating genomic analyses due to its well-characterized genome. In our recent study, We established an expression benchmark and a splicing junction benchmark using publicly available RNA-seq data of GM24385. At the expression level, the benchmark comprises 8,991 genes positively expressed and 1,296 genes negatively expressed. At the splicing level, the benchmark includes 38,110 positive junctions and 4,195 negative junctions. Sensitivity is measured by the detection rate of positively identified genes or junctions, while specificity is measured by the detection rate of negatively identified genes or junctions.

### Tissue-specific gene and isoform expression

The expression profile of each tissue was obtained from Gtex database. The tissue preference score (Pscore) of a gene/isoform is defined as:

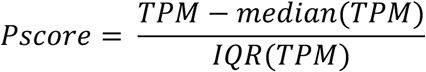

Where TPM is the TPM of the gene/isoform in a given tissue; median (TPM) is the median TPM of this gene/isoform across all Gtex tissues; IQR (TPM) is the interquartile range of TPM across all Gtex tissues. A gene/isoform with Pscore ≥ 2 in a tissue was defined to have specific expression to this tissue ^22^.

### RNA outlier analysis

We calculated the P-value of *ITPR1* splicing change using FRASER v1.99.1 ^23^. A pool of 26 deep RNA-seq on fibroblast was used as the reference data. Total reads were downsampled to 1000M, 200M, and 50M to investigate the impact of sequencing depth on the significance of the outlier.

Power analysis of expression outlier was performed using OUTRIDER v1.17.2 ^24^. Theta values of all genes were calculated based on a dataset of 73 regular-depth RNA-seq on fibroblasts ^10^.

### Minimum required sequencing depth (MRSD)

MRSD is specific to each sample type and was calculated using the same logic as previously described ^16^. The junction-level MRSD of a gene depends on the desired proportion of its splice junctions (*p*) to be covered by a desired number of sequencing reads (*r*). We used the median (junction read count)/(total read count) ratio across three deep RNA-seq data for each CAT as the baseline expression abundance of the junction (*a*). Then MRSD was calculated as:

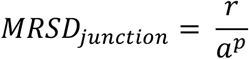

Where *a^p^* is the expression abundance of the junction at the p quantile of all junctions in the gene, and *r* is the desired number of sequencing reads. A junction is regarded as unfeasible if the median junction read count is 0. MRSD was calculated under different desired junction read counts (10, 20, 50, and 100) and different proportions of junctions (75% and 95%) covered for each gene. The expression-level MRSD of a gene only depends on the desired number of sequencing reads (*r*). We used the median (gene read count)/(total read) count ratio across three deep RNAseq data for each CAT as the baseline expression abundance of the gene (*a*). Then MRSD was calculated as:

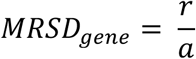

A gene is regarded as unfeasible if the median read count is 0. MRSD was calculated under different desired read counts (500, 1450, and 2000).

For the power analysis of gene-based outlier analysis, a cohort of 119 RNA-seq samples from a previous study was used. Power analysis was performed using Outrider v1.17.2 based on a negative binomial distribution of the read counts ^24^. As a cutoff for power analysis, we selected the read count required to detect a 50% percent downregulation at a genome-wide significance (P value=0.05/19217).

### Naturally occurring splicing variations

Based on the splicing matrices from 18 fibroblast deep RNA-seq data and 18 regular-depth RNA-seq data from our recent study^10^, we constructed two splicing variation databases using a strategy similar to that previously reported ^18^. We merged STAR splicing junction matrices for each dataset. Splicing junctions were filtered to those that span one annotated splice site and one unannotated splice site. For each unannotated junction, the number of occurrences among the 18 deep RNA-seq data was tallied and grouped by the canonical splice sites where they occurred. At each canonical splice donor/acceptor site, unannotated splicing junctions were ranked based on their occurrence in the 18 reference samples We collected 28 genetic variants with known splicing consequences from our in-house database and the previous study ^18^. For each variant, we predicted the splicing consequences using our deep RNA-seq database and the original SpliceVault. The SpliceVault data was filtered for fibroblast samples (n = 520) for comparison. For the consequence(s) of each variant, their ranks from our deep RNA-seq database and SpliceVault were recorded and compared.

## Data and code availability

Raw RNA-seq data from this study have been deposited in the NCBI Sequence Read Archive (SRA) under accession number PRJNA1150958.

## Data Availability

https://www.ncbi.nlm.nih.gov/bioproject/PRJNA1150958

## Acknowledgments

Research reported in this publication was supported by the National Institute Of Neurological Disorders And Stroke of the National Institutes of Health (U01HG007709 and U01HG007942) and the grant from National Human Genome Research Institute (R35HG011311). The content is solely the responsibility of the authors and does not necessarily represent the official views of the National Institutes of Health.

## References

1 Linderman, M. D. et al. Analytical validation of whole exome and whole genome sequencing for clinical applications. BMC Med Genomics 7, 20 (2014). 10.1186/1755-8794-7-20

2 Rehm, H. L. et al. ACMG clinical laboratory standards for next-generation sequencing. Genet Med 15, 733–747 (2013). 10.1038/gim.2013.92

3 Marshall, C. R. et al. Best practices for the analytical validation of clinical whole-genome sequencing intended for the diagnosis of germline disease. NPJ Genom Med 5, 47 (2020). 10.1038/s41525-020-00154-9

4 External, R. N. A. C. C. Proposed methods for testing and selecting the ERCC external RNA controls. BMC Genomics 6, 150 (2005). 10.1186/1471-2164-6-150

5. Consortium, E. P. A user’s guide to the encyclopedia of DNA elements (ENCODE). PLoS Biol 9, e1001046 (2011). 10.1371/journal.pbio.1001046

6 Fresard, L. et al. Identification of rare-disease genes using blood transcriptome sequencing and large control cohorts. Nat Med 25, 911–919 (2019). 10.1038/s41591-019-0457-8

7 Murdock, D. R. et al. Transcriptome-directed analysis for Mendelian disease diagnosis overcomes limitations of conventional genomic testing. J Clin Invest 131 (2021). 10.1172/JCI141500

8 Kremer, L. S. et al. Genetic diagnosis of Mendelian disorders via RNA sequencing. Nat Commun 8, 15824 (2017). 10.1038/ncomms15824

9 Aicher, J. K., Jewell, P., Vaquero-Garcia, J., Barash, Y. & Bhoj, E. J. Mapping RNA splicing variations in clinically accessible and nonaccessible issues to facilitate Mendelian disease diagnosis using RNA-seq. Genet Med 22, 1181–1190 (2020). 10.1038/s41436-020-0780-y

10 Zhao, S. et al. Clinical validation of RNA sequencing for Mendelian disorder diagnostics. medRxiv, 2024.2008.2015.24312057 (2024). 10.1101/2024.08.15.24312057

11 Chin, M. H. et al. Induced pluripotent stem cells and embryonic stem cells are distinguished by gene expression signatures. Cell Stem Cell 5, 111–123 (2009). 10.1016/j.stem.2009.06.008

12 O’Neil, D., Glowatz, H. & Schlumpberger, M. Ribosomal RNA depletion for efficient use of RNA-seq capacity. Curr Protoc Mol Biol **Chapter** 4, Unit 4 19 (2013). 10.1002/0471142727.mb0419s103

13 Keehan, L. et al. A novel de novo intronic variant in ITPR1 causes Gillespie syndrome. Am J Med Genet A 185, 2315–2324 (2021). 10.1002/ajmg.a.62232

14 Li, S. et al. The clinical utility and diagnostic implementation of human subject cell transdifferentiation followed by RNA sequencing. Am J Hum Genet 111, 841–862 (2024). 10.1016/j.ajhg.2024.03.007

15 Terkelsen, T. et al. CRISPR activation to characterize splice-altering variants in easily accessible cells. Am J Hum Genet 111, 309–322 (2024). 10.1016/j.ajhg.2023.12.024

16 Rowlands, C. F. et al. MRSD: A quantitative approach for assessing suitability of RNA-seq in the investigation of mis-splicing in Mendelian disease. Am J Hum Genet 109, 210–222 (2022). 10.1016/j.ajhg.2021.12.014

17 Park, E., Pan, Z., Zhang, Z., Lin, L. & Xing, Y. The Expanding Landscape of Alternative Splicing Variation in Human Populations. Am J Hum Genet 102, 11–26 (2018). 10.1016/j.ajhg.2017.11.002

18 Dawes, R. et al. SpliceVault predicts the precise nature of variant-associated mis-splicing. Nat Genet 55, 324–332 (2023). 10.1038/s41588-022-01293-8

19 Qi, T. et al. Genetic control of RNA splicing and its distinct role in complex trait variation. Nat Genet 54, 1355–1363 (2022). 10.1038/s41588-022-01154-4

20 Halvardson, J., Zaghlool, A. & Feuk, L. Exome RNA sequencing reveals rare and novel alternative transcripts. Nucleic Acids Res 41, e6 (2013). 10.1093/nar/gks816

21. Nicolas-Martinez, E. C., et al. RNA variant assessment using transactivation and transdifferentiation. Am J Hum Genet 111, 1673-1699 (2024). 10.1016/j.ajhg.2024.06.018

22 Sonawane, A. R. et al. Understanding Tissue-Specific Gene Regulation. Cell Rep 21, 1077–1088 (2017). 10.1016/j.celrep.2017.10.001

23 Scheller, I. F., Lutz, K., Mertes, C., Yepez, V. A. & Gagneur, J. Improved detection of aberrant splicing with FRASER 2.0 and the intron Jaccard index. Am J Hum Genet 110, 2056–2067 (2023). 10.1016/j.ajhg.2023.10.014

24 Brechtmann, F. et al. OUTRIDER: A Statistical Method for Detecting Aberrantly Expressed Genes in RNA Sequencing Data. Am J Hum Genet 103, 907–917 (2018). 10.1016/j.ajhg.2018.10.025

