## Supplementary* for "The Utility of Ultra-Deep RNA sequencing in Mendelian Disorder Diagnostics"

### Supplementary information

#### Supplementary Figures

##### Figure S1. Illumina and Ultima sequencing platform comparison

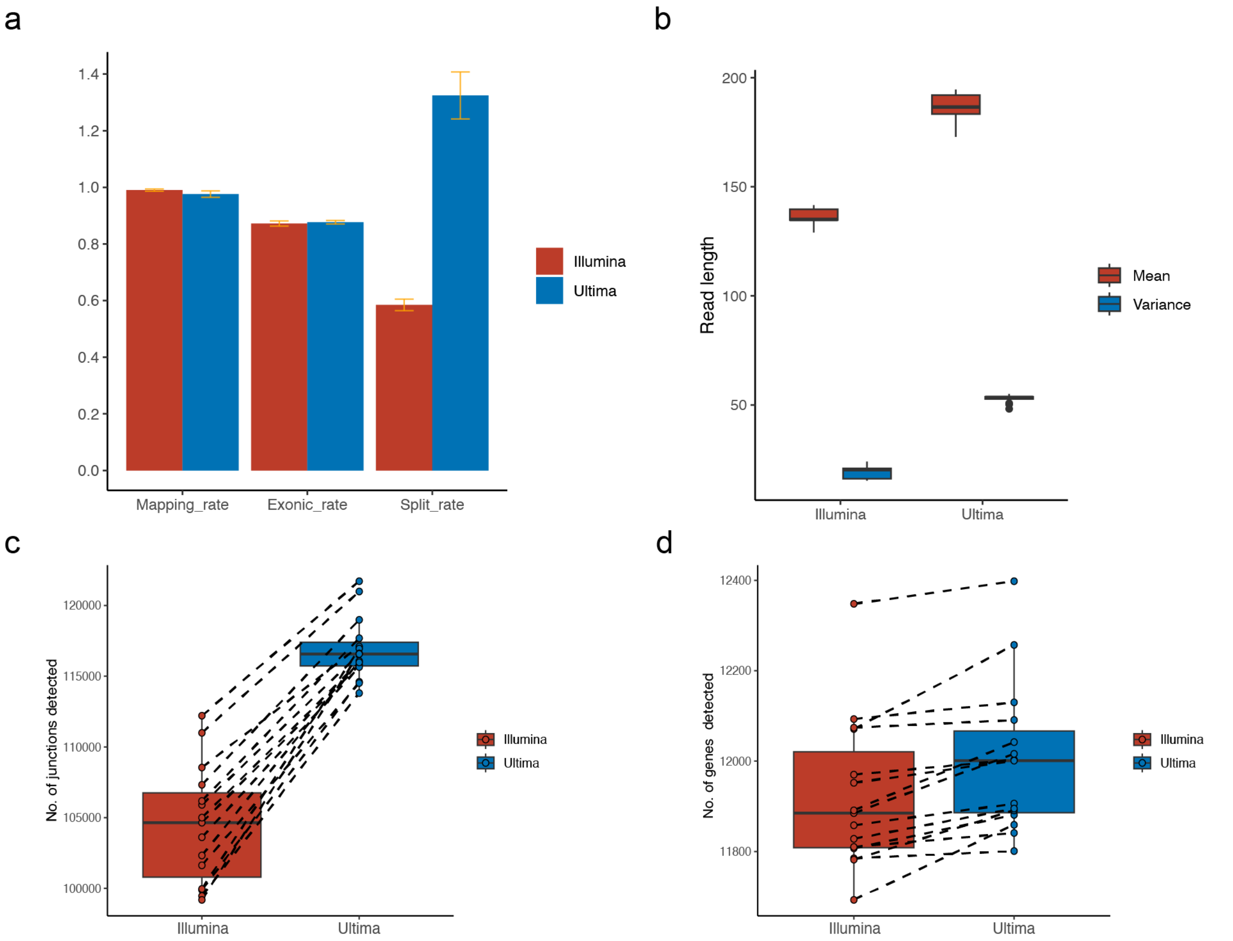


We compared the quality metrics of Ultima RNA-seq against Illumina RNA-seq using 15 fibroblast samples sequenced using both techniques. Each pair of Ultima/Illumina data was downsampled to the same number of total reads. a, Ultima and Illumina data had similar mapping rates and exonic rate. b and c, Ultima sequencing reads showed a higher split rate due to their longer read lengths, which resulted in a higher number of splicing junctions detected. d, The no. of genes detected in Ultima and Illumina sequencing.

### Figure S2. Correlation of gene read count between Ultima and Illumina sequencing

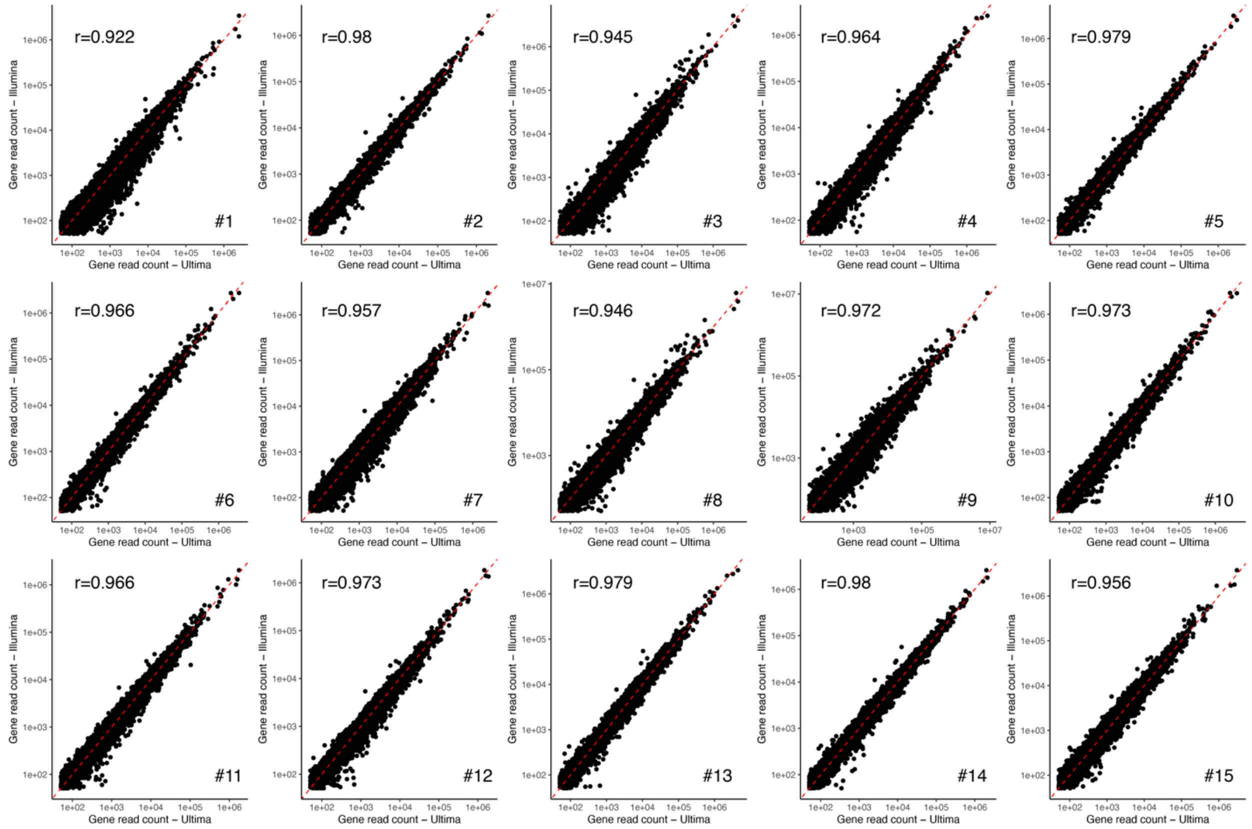


Read count of each gene and Pearson r coefficient between each pair of Ultima/Illumina data is shown.

#### Figure S3. Library design of Ultima sequencing

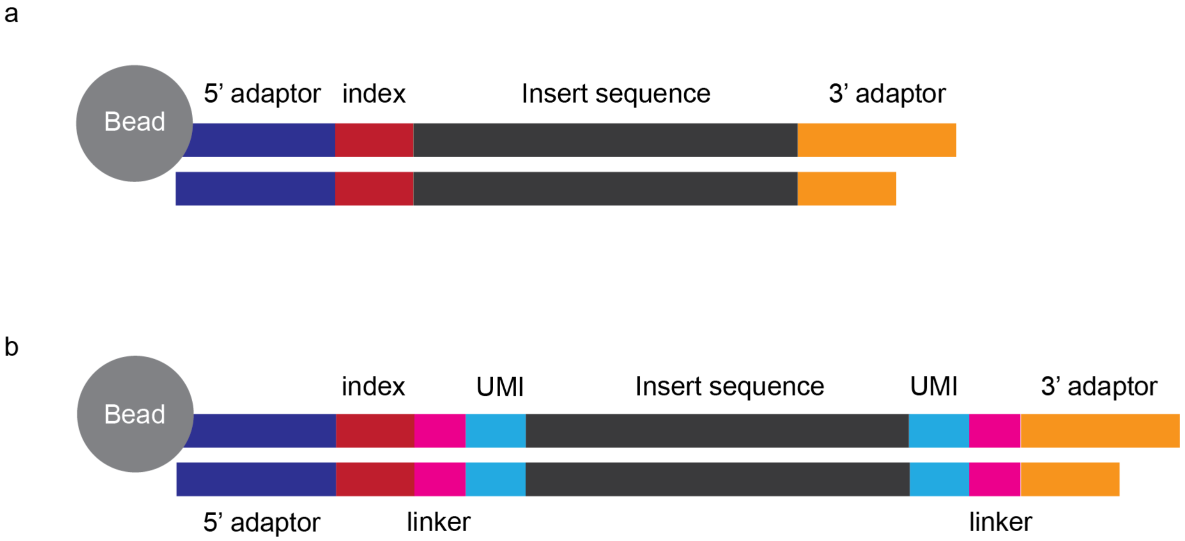


A, the standard library design of Ultima sequencing. B, Ultima library design with unique molecular identifiers (UMI).

#### Figure S4. Exonic rate at different sequencing depth

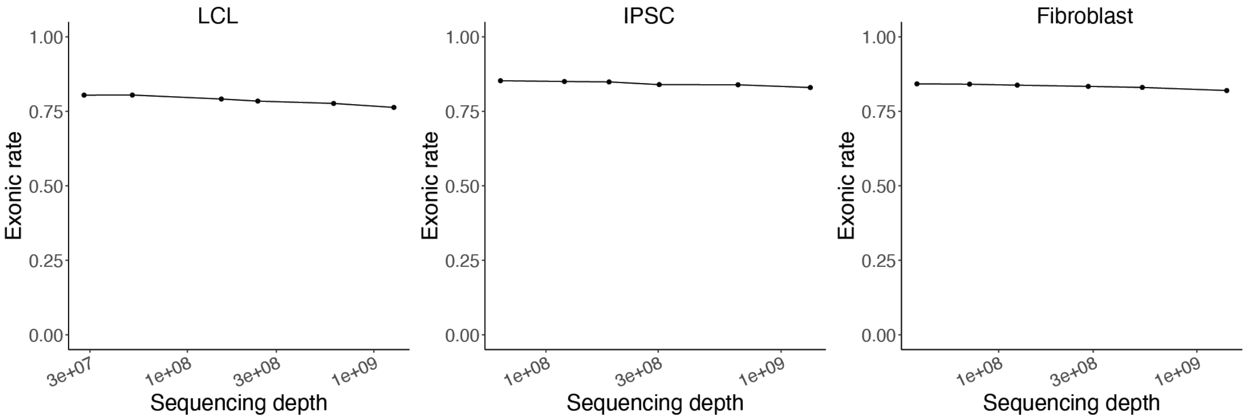


The overall exonic rate of three samples sequenced at different sequencing depth. The samples are GM12878 (LCL), BG2111P0002 (IPSC), and UD1902P0001 (fibroblast).

#### Figure S5. The relation between the junction ratio and the expression count

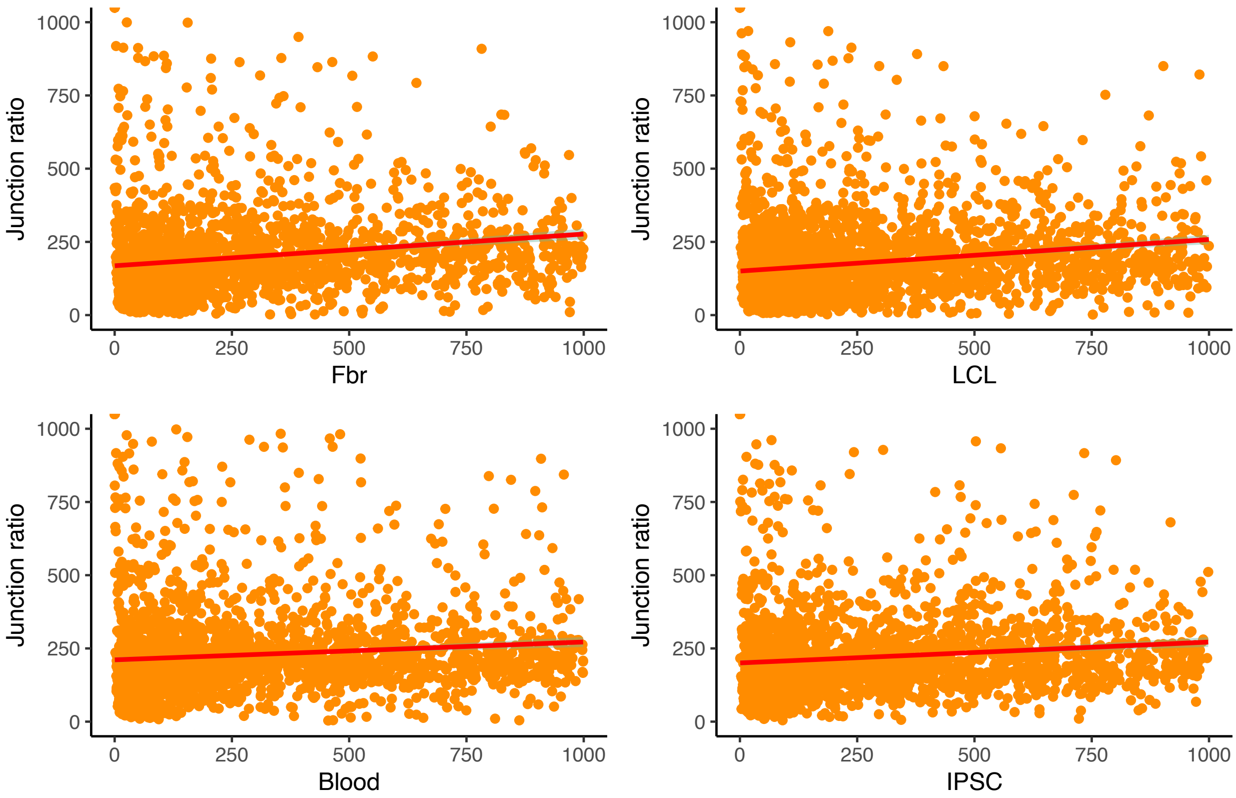


For each gene of each sample type, we used the deep RNA-seq data to calculate the ratio of junction-spanning reads versus total reads and used this metric to evaluate the reliability of gene expression. Only genes with a read count < 1000 are shown because most false positive expressions are at the low-expression end.

Figure S6. Distribution of gene read count across clinically accessible tissues

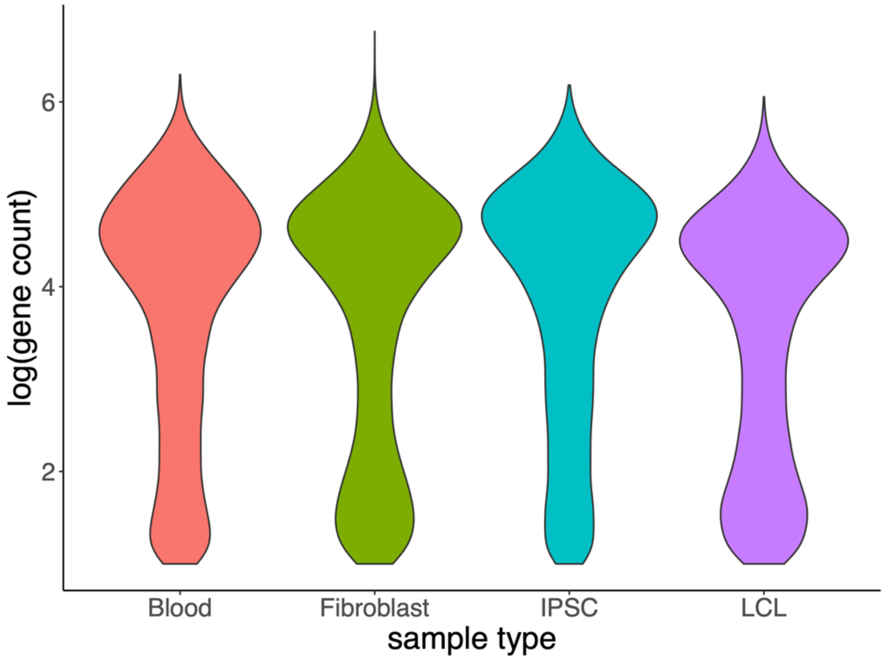


LCL, lymphoblastoid cells; IPSC, induced pluripotent stem cells.

#### Figure S7. Low-expressing genes in clinically accessible tissues

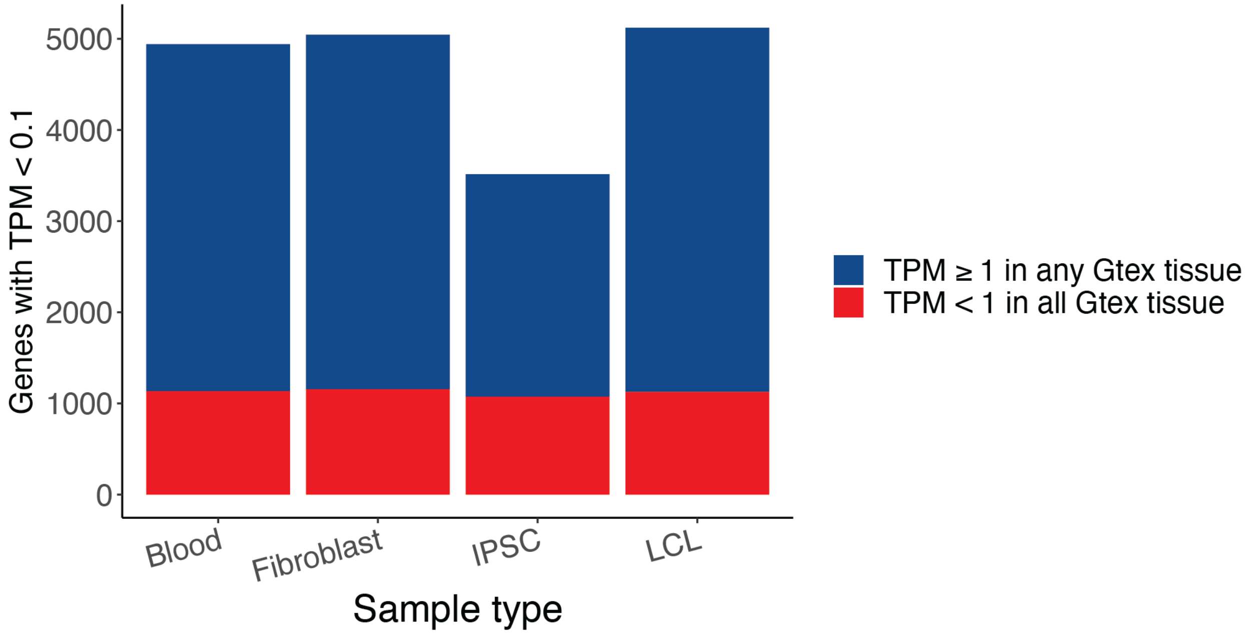


The proportion of low-expressing genes in clinically accessible tissues that are high expressing in at least one Gtex tissue or low-expressing in all Gtex tissue.

#### Figure S8. Tissue-specific gene expression in Gtex

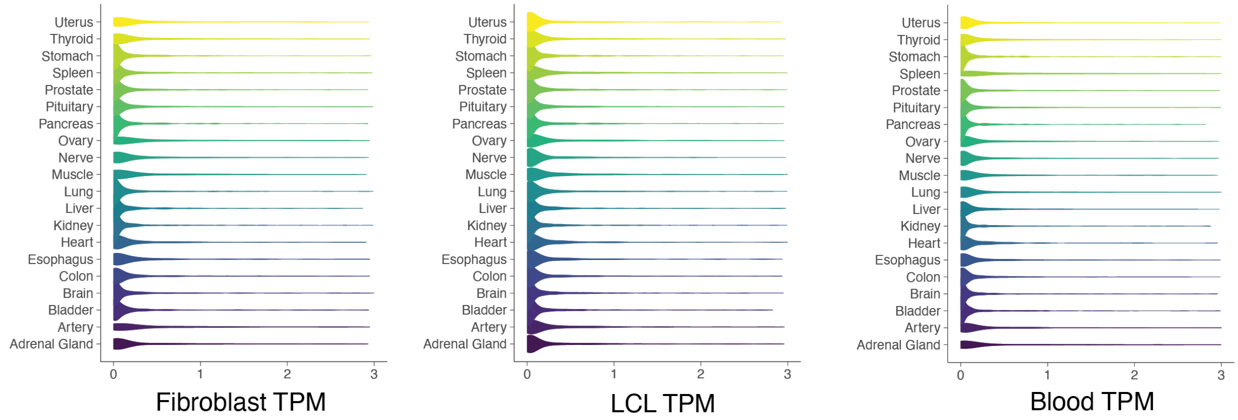


Tissue-specific genes (tissue preference score > 2) in Gtex showed a depletion of expression in CATs (fibroblast, blood, and LCL).

#### Figure S9. Leave-one-out experiment on high-expressing genes

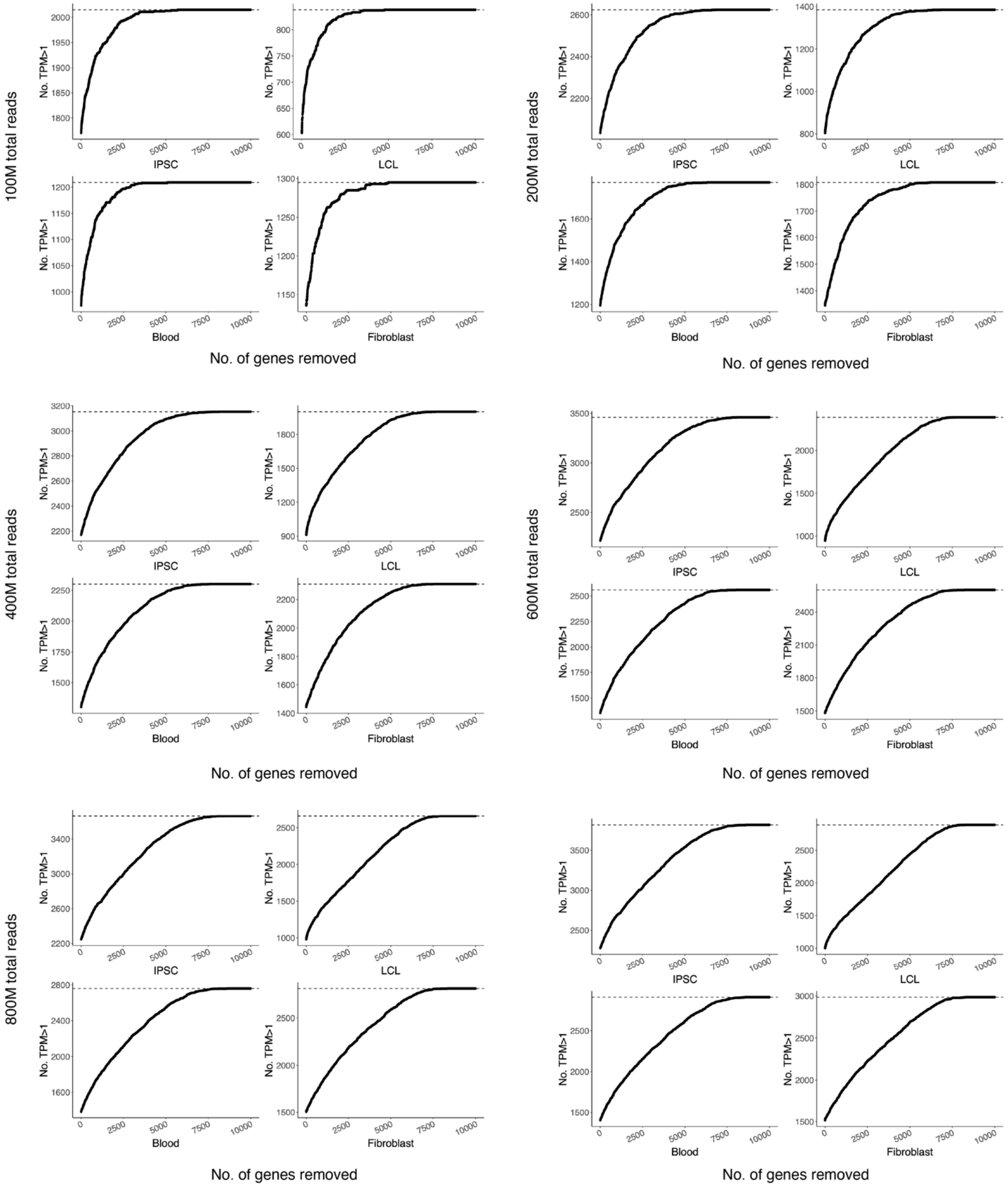


We performed in silico removal of high-expressing genes one by one and evaluated the number of additional genes whose TPM exceeded one. We performed this leave-one-out experiment at different sequencing depths.

#### Figure S11. Correlation between our MRSD data and the original MRSD

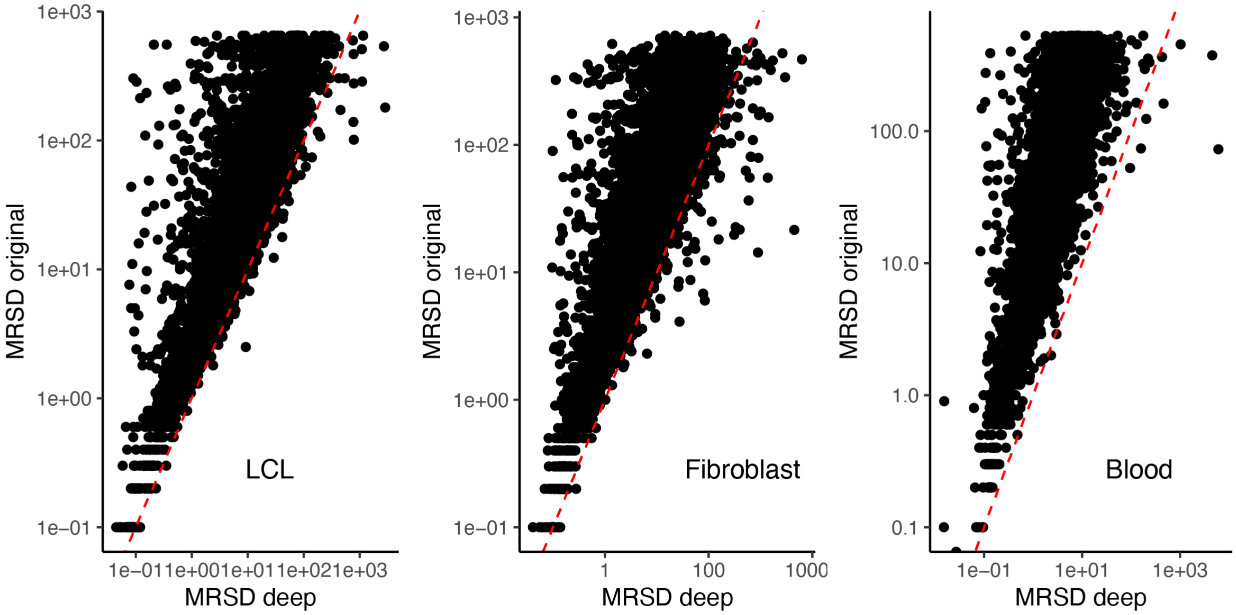


Correlation of MRSD metrics between our data and the original MRSD data. MRSD was calculated at a required junction coverage of 95% and junction read of 10.

#### Figure S10. Distribution of theta value in expression outlier analysis

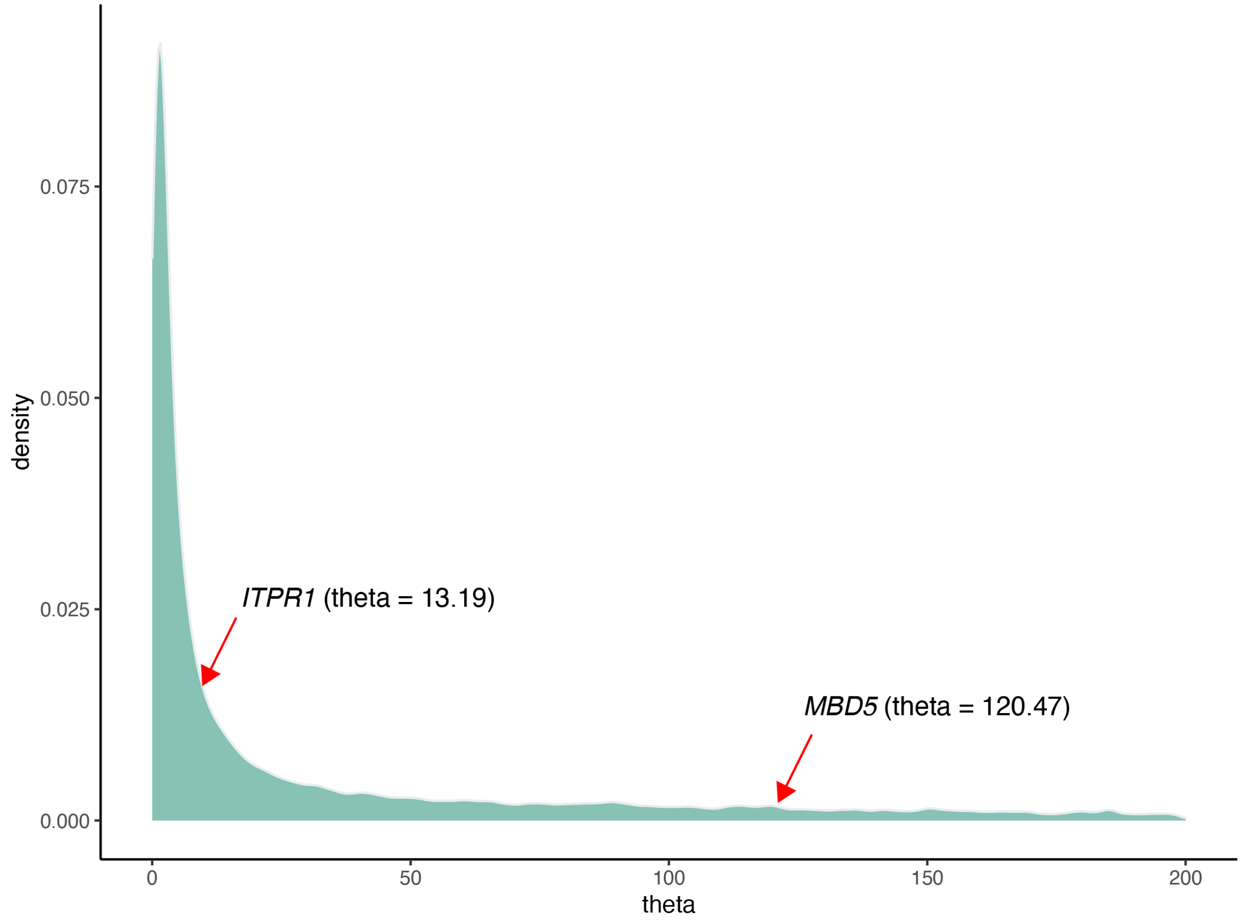


The theta value indicates the level of variation of a gene in a negative binomial distribution, which was used for the expression outlier analysis. Here we plot the distribution of theta values that were calculated based on a dataset of 73 regular-depth RNA-seq on fibroblasts [13].

### Supplementary Tables

#### Table S1. Sequencing design

| SampleID | Sample type | Group | Source | Sequence design |
|---|---|---|---|---|
| **UD2210P0003** | Fibroblast | Validation | UDN | Ultima and Illumina |
| **UD2210P0004** | Fibroblast | Validation | UDN | Ultima and Illumina |
| **UD1708P0003** | Fibroblast | Validation | UDN | Ultima and Illumina |
| **UD2211P0001** | Fibroblast | Validation | UDN | Ultima and Illumina |
| **UD2210P0005** | Fibroblast | Validation | UDN | Ultima and Illumina |
| **UD1712P0002** | Fibroblast | Validation | UDN | Ultima and Illumina |
| **UD1911P0001** | Fibroblast | Validation | UDN | Ultima and Illumina |
| **UD1708P0002** | Fibroblast | Validation | UDN | Ultima and Illumina |
| **UD2010P0002** | Fibroblast | Validation | UDN | Ultima and Illumina |
| **UD1509P0001** | Fibroblast | Validation | UDN | Ultima and Illumina |
| **UD1710P0001** | Fibroblast | Validation | UDN | Ultima and Illumina |
| **UD2209P0003** | Fibroblast | Validation | UDN | Ultima and Illumina |
| **UD2210P0007** | Fibroblast | Validation | UDN | Ultima and Illumina |
| **UD2210P0008** | Fibroblast | Validation | UDN | Ultima and Illumina |
| **UD2210P0001** | Fibroblast | Validation | UDN | Ultima and Illumina |
| **GM24385** | LCL | Validation | UDN | Ultima RNA-seq at different depth |
| **GM24385** | LCL | Main | Coriell | Ultima deep RNA-seq and downsample |
| **GM12878** | LCL | Main | Coriell | Ultima deep RNA-seq and downsample |
| **GM12877** | LCL | Main | Coriell | Ultima deep RNA-seq and downsample |
| **BG2111P0002** | IPSC | Main | In-house | Ultima deep RNA-seq and downsample |
| **BG2111P0003** | IPSC | Main | In-house | Ultima deep RNA-seq and downsample |
| **GM27730** | IPSC | Main | Coriell | Ultima deep RNA-seq and downsample |
| **UD1902P0001** | Fibroblast | Main | UDN | Ultima deep RNA-seq and downsample |
| **UD2008P0001** | Fibroblast | Main | UDN | Ultima deep RNA-seq and downsample |
| **BG2302P0023** | Fibroblast | Main | UDN | Ultima deep RNA-seq and downsample |
| **BG2305P0017** | Blood | Main | In-house | Ultima deep RNA-seq and downsample |
| **BG2305P0014** | Blood | Main | In-house | Ultima deep RNA-seq and downsample |
| **BG2305P0015** | Blood | Main | In-house | Ultima deep RNA-seq and downsample |
| **UD1705P0001** | Fibroblast | Splice Vault | UDN | Ultima deep RNA-seq |
| **UD2008P0002** | Fibroblast | Splice Vault | UDN | Ultima deep RNA-seq |
| **UD2010P0007** | Fibroblast | Splice Vault | UDN | Ultima deep RNA-seq |
| **UD2101P0001** | Fibroblast | Splice Vault | UDN | Ultima deep RNA-seq |
| **UD1909P0001** | Fibroblast | Splice Vault | UDN | Ultima deep RNA-seq |
| **UD2011P0001** | Fibroblast | Splice Vault | UDN | Ultima deep RNA-seq |
| **UD2312P0029** | Fibroblast | Splice Vault | UDN | Ultima deep RNA-seq |

| | | | | |
|---|---|---|---|---|
| **UD2312P0030** | Fibroblast | Splice Vault | UDN | Ultima deep RNA-seq |
| **UD2312P0031** | Fibroblast | Splice Vault | UDN | Ultima deep RNA-seq |
| **UD2312P0032** | Fibroblast | Splice Vault | UDN | Ultima deep RNA-seq |
| **UD2312P0033** | Fibroblast | Splice Vault | UDN | Ultima deep RNA-seq |
| **UD2312P0034** | Fibroblast | Splice Vault | UDN | Ultima deep RNA-seq |
| **UD2312P0035** | Fibroblast | Splice Vault | UDN | Ultima deep RNA-seq |
| **UD1702P0001** | Fibroblast | Splice Vault | UDN | Ultima deep RNA-seq |
| **UD1705P0002** | Fibroblast | Splice Vault | UDN | Ultima deep RNA-seq |

Abbreviations: UDN, Undiagnosed Diseases Network; LCL, lymphoblastoid cells; IPSC, induced pluripotent stem cells.

#### Table S2. Benchmarking of Ultima RNA-seq at different sequencing depths

| sampleID | total_reads | Expression sensitivity | Expression specificity | Junction sensitivity | Junction specificity |
|---|---|---|---|---|---|
| **Ultima-R1** | 39495128 | 0.9938 (0.9919-0.9952) | 1 (0.997-1) | 0.9525 (0.9503-0.9546) | 0.9988 (0.9972-0.9995) |
| **Ultima-R2** | 73931437 | 0.9977 (0.9964-0.9985) | 1 (0.997-1) | 0.9833 (0.982-0.9846) | 0.9952 (0.9926-0.9969) |
| **Ultima-R3** | 137456463 | 0.999 (0.9981-0.9995) | 0.9977 (0.9932-0.9992) | 0.9957 (0.995-0.9964) | 0.9955 (0.9929-0.9971) |
| **Ultima-R4** | 229833559 | 0.9996 (0.9989-0.9998) | 0.9969 (0.9921-0.9988) | 0.999 (0.9987-0.9993) | 0.9952 (0.9926-0.9969) |
| **Ultima-R5** | 538090163 | 1 (0.9996-1) | 0.9931 (0.9869-0.9963) | 0.9999 (0.9998-1) | 0.9914 (0.9881-0.9938) |
| **Ultima-R6** | 825795035 | 1 (0.9996-1) | 0.9853 (0.9772-0.9906) | 1 (0.9999-1) | 0.9878 (0.9841-0.9907) |
| **Illumina-R1** | 148912894 | 0.9993 (0.9985-0.9997) | 1 (0.997-1) | 0.9967 (0.9961-0.9972) | 0.999 (0.9976-0.9996) |

In our recent study, we established provisional expression and splicing benchmarks using short-read and long-read RNA-seq data of GM24385 (HG002) lymphoblastoid sample from the Genome in a Bottle Consortium (med achieve). The benchmark data are based on short-read and long-read sequencing data from independent laboratories. At the expression level, the benchmark comprises 8,991 genes positively expressed and 1,296 genes negatively expressed. At the splicing level, the benchmark includes 38,110 positive junctions and 4,195 negative junctions. Sensitivity is measured by the detection rate of positively identified genes or junctions, while specificity is measured by the detection rate of negatively identified genes or junctions. Here we present the benchmarking result of six Ultima runs at different sequencing depths and an Illumina run at a sequencing depth of around 150M reads.

#### Table S3. Sequencing depth and duplication rate of deep RNA-seq

| SampleID | Sample type | Total reads | Total reads after dedup | Duplication rate | Corrected duplication rate |
|---|---|---|---|---|---|
| GM24385 | LCL | 2551462649 | 1363885198 | 0.51119641 | 0.17004607 |
| GM12878 | LCL | 3505224253 | 1280686414 | 0.36536504 | 0.37719471 |
| GM12877 | LCL | 2481670473 | 1286309415 | 0.49630999 | 0.20877628 |
| BG2111P0002 | IPSC | 2256319282 | 1331443203 | 0.59009521 | 0.14273151 |
| BG2111P0003 | IPSC | 2827589890 | 1726235124 | 0.61049699 | 0.1561387 |
| GM27730 | IPSC | 2494484053 | 1546995400 | 0.62016648 | 0.12973232 |
| UD1902P0001 | Fibroblast | 3068749214 | 1414947891 | 0.46108293 | 0.21518137 |
| UD2008P0001 | Fibroblast | 3274256453 | 1551775734 | 0.47393225 | 0.20074413 |
| BG2302P0023 | Fibroblast | 2771719302 | 1402160808 | 0.46967527 | 0.27930863 |
| BG2305P0017 | Blood | 3726592540 | 1414521526 | 0.37957504 | 0.34686539 |
| BG2305P0014 | Blood | 3316482334 | 1357710573 | 0.40938272 | 0.25162487 |
| BG2305P0015 | Blood | 3759418918 | 1348749129 | 0.35876532 | 0.36901384 |

The corrected duplication rate was calculated using medium-expressing genes (100 < read count < 10000).

#### Table S4. Correlations between in vitro and in silico downsample

| patient_ID | SequenceID | Sample type | Total reads | Correlation | No. of expressed gene-in vitro | No. of expressed gene-in silico |
|---|---|---|---|---|---|---|
| GM12878 | R1 | LCL | 27909716 | 0.997 | 10311 | 10700 |
| GM12878 | R2 | LCL | 50660918 | 0.998 | 10937 | 11212 |
| GM12878 | R3 | LCL | 152082113 | 0.999 | 11977 | 12186 |
| GM12878 | R4 | LCL | 238510474 | 0.999 | 12495 | 12658 |
| GM12878 | R5 | LCL | 608510230 | 0.999 | 13656 | 13790 |
| BG2111P0002 | R1 | IPSC | 64025647 | 0.968 | 13012 | 12372 |
| BG2111P0002 | R2 | IPSC | 119775817 | 0.973 | 13654 | 13066 |
| BG2111P0002 | R3 | IPSC | 185439721 | 0.976 | 14073 | 13775 |
| BG2111P0002 | R4 | IPSC | 303103580 | 0.978 | 14618 | 14431 |
| BG2111P0002 | R5 | IPSC | 655714575 | 0.98 | 15341 | 15071 |
| UD1902P0001 | R1 | Fibroblast | 38723274 | 0.997 | 11271 | 11609 |
| UD1902P0002 | R2 | Fibroblast | 71379350 | 0.998 | 11780 | 12052 |
| UD1902P0003 | R3 | Fibroblast | 124212876 | 0.998 | 12202 | 12465 |
| UD1902P0004 | R4 | Fibroblast | 283210981 | 0.999 | 12961 | 13080 |
| UD1902P0005 | R5 | Fibroblast | 530489447 | 0.999 | 13411 | 13528 |

Each of the three samples was sequenced at different depths (R1-R5). The data at the highest depth was downsampled to the same depth as the data at lower sequencing depths. We compared the correlation of expression profiles (Pearson coefficient of read count and no. of expressed genes) between in vitro and in silico downsampled data.

### Supplementary Data

#### Data S1. The relation between the sequencing depth of RNA-seq and number of genes detected

#### Data S2. The relation between proportion of tissue-specific genes detected and sequencing depth.

Tissue-specific genes were derived from the Gtex database.

#### Data S3. The relation between the sequencing depth of RNA-seq and number of isoforms detected

#### Data S4. The relation between the proportion of tissue-specific isoforms detected and sequencing depth.

Tissue-specific isoforms were derived from the Gtex database.

#### Data S5. Junction level minimum required depth

Minimum required depth (MRSD) under different targeted junction coverage and percent of junctions covered for each gene.

#### Data S6. Expression level minimum required depth

Minimum required depth (MRSD) under different targeted gene coverage.

#### Data S7. Splicing variation databased constructed using deep RNA-seq data

Based on the splicing matrices from 18 fibroblast deep RNA-seq data, we constructed a splicing variation database.

#### Data S8. Positive variants with known splicing consequences
